# Patient Needs and Requirements for a Decision Aid Navigator System (DEAN) Patient Portal: A Qualitative Study

**DOI:** 10.64898/2026.10.12.26365343

**Authors:** Brad Morse, Carrie Reale, An T. Nguyen, Erin Latella, Hannah Bauguess, Pamela Roberts, Spencer L SooHoo, Robert El-Kareh, Janelle Faiman, Andrey Soares, Lisa M. Schilling

**Affiliations:** Division of General Internal Medicine, University of Colorado Anschutz, Aurora, CO, USA; Adult and Child Center for Outcomes Research and Delivery Science, University of Colorado School of Medicine, Aurora, CO, USA; Center for Research and Innovation in Systems Science, Vanderbilt University Medical Center, Nashville. TN, USA; Department of Anesthesiology, Vanderbilt University Medical Center, Nashville, TN, USA; Department of Medical Affairs and Department of Physical Medicineand Rehabilitation, Cedars-Sinai Medical Center, Los Angeles, CA, USA; Department of Computational Biomedicine, Division of Informatics, Cedars-Sinai Medical Center, Los Angeles, CA, USA; Division of Biomedical Informatics, University of California, San Diego, La Jolla, CA, USA

**Author notes:** **Corresponding author:** Brad Morse, PhD, MA, University of Colorado Anschutz, School of Medicine 1890 N Revere Ct Ste: P12-3221, Aurora, CO 80045, USA.

**Keywords:** Decision aid, shared decision-making, patient portal, focus groups, qualitative research

## Abstract

**Background:** Patient Decision aids (PtDAs) are important tools that support shared decision-making (SDM) between clinicians and patients, enabling patients to be more informed and engaged in decisions regarding their care. The use of PtDAs can increase patient knowledge, reduce decisional regret, and engage the clinician and patient in meaningful dialogue. Shared decision-making relies on decision aids, but how patients receive the PtDA can vary. This qualitative study elucidated patient familiarity with PtDAs and the barriers, needs, and requirements of patients to access and confidently use DAs, including when received via a clinician-facing EHR-integrated Decision Aid Navigator System (DEANS).

**Methods:** We conducted six focus groups (FG) with participants who were recruited from two academic medical centers. Focus groups included an introduction to what a PtDA is and an exploration of how patients or family members, i.e., people who had not made a medical decision for themselves using a PtDA, but who have helped a family member, had previously used PtDAs in their healthcare. The FGs focused on PtDA-users, affordances and barriers to using PtDAs, and PtDA-user needs and requirements of a proposed DEAN patient portal. We applied Rapid Qualitative Analysis (RQA) to summarize the data, using a consensus making approach to identify emerging themes.

**Results:** We identified three overarching themes: 1) Patients had varied familiarity with PtDAs and often confused them with general health tools, but found value in SDM, 2) Patients perceived technology accessibility, PtDA credibility, and time as barriers potentially impeding PtDA use, and 3) Patients valued flexible, user-friendly, and accessible PtDAs that support patient-led decisions, particularly when introduced by clinicians and delivered via a patient portal.

**Conclusion:** This study provides patient-derived insights into factors that influence PtDA uptake, highlighting common misconceptions about PtDAs and identifying key patient priorities such as accessibility, flexibility, clear content, and clinician-guided usage. Patient also expected clinicians to be familiar with PtDAs, especially when the PtDA covers a complex decision. These findings can inform the development of more patient-centered SDM infrastructure and strategies to support PtDA use in clinical practice, including clinician-supported delivery, accessibility, and integration into routine care to strengthen shared decision-making.

## Background

A patient decision aid (PtDA) is a tool designed and developed to support patients, proxies, and family members to make informed and intentional decisions.^1^ PtDAs are often used in the context of shared decision-making (SDM) discussion between patients and clinicians.^2–4^ PtDAs can take various forms, including PDFs, videos, or web-based tools^5^, and can facilitate meaningful conversations between patients and clinicians. Most PtDA are used as part of a SDM discussion, whether the patient uses the PtDA first, asynchronously in preparation for a discussion with a clinician, or whether the PtDA is used synchronously during a patient/caregiver-clinician encounter. PtDAs support autonomy and help ensure that medical decisions align with what matters most to the individual.^2^ Research has shown that PtDAs increase patient involvement and satisfaction with the SDM process.^6–8^

Treatment options are presented in PtDAs by providing evidence-based information about the benefits, risks, and potential outcomes of each choice. PtDAs are particularly useful in situations where there is no single “best” option, and the right decision depends on the patient’s values and preferences. PtDAs aim to clarify personal values, improve knowledge, and reduce decisional conflict. They are especially valuable in preference-sensitive decisions, such as choosing between surgical and non-surgical treatments or deciding on cancer screening options. Other characteristics that make a medical decision preference-sensitive include a state of no clinical consensus^9^ and multiple valid treatment options and tradeoffs,^10^ i.e., quality and anticipated length of patient’s life. For example, a PtDA might help a patient with knee osteoarthritis decide between physical therapy, medication, or joint replacement surgery by presenting comparative data and prompting reflection on lifestyle and goals.^11,12^

As healthcare becomes more personalized, PtDAs are increasingly recognized as essential components of ethical and patient-centered care.^12^ Their use has been linked to improved health outcomes, reduced unnecessary interventions, and greater alignment between patient choices and clinical recommendations.^13,14^ As such, PtDAs represent a critical advancement in promoting informed, value-congruent healthcare.^2^ The optimal use of PtDAs is complicated by the nature of the decision itself. Simple decisions may be more amenable to independent patient engagement, while perceived complex decisions often require nuanced clinical guidance.^15^ Prior studies have explored the timing and association of PtDAs to clinical visits, with some trials assessing the impact of providing PtDAs pre-visit or entirely outside the clinical encounter.

Evidence suggests that the effectiveness of PtDAs may depend when and how often the patient receives the decision aid. Pre-visit delivery has been shown to improve preparedness, but not consistently producing improved decision quality or clinical outcomes.^12,16^ The impact a PtDA has on decision-making, related to the timing of a patient’s receipt of PtDA, is not settled. Some clinicians prefer that their patients receive PtDAs before a visit due to time constraints and delivery of the PtDA,^17–20^ while other clinicians believe in-visit receipt of the PtDA supports better SDM.^21,22^

### Barriers facing patient decision aid uptake

Despite the proven benefits of PtDA use,^23,24^ several barriers continue to prevent use. Patient-specific barriers include poor health literacy, which can hinder engagement in SDM due to difficulty understanding complex medical information and jargon.^25,26^ Emotional distress is another factor; the pressure of making a serious and potentially final decision can provoke fear and anxiety, reducing a patient’s ability to use PtDAs effectively to engage in a SDM process.^27^ General distrust in the healthcare system, often rooted in historical injustices^28,29^, can make patients hesitant to engage with tools perceived as part of that system.^27,30^ Patients can feel overwhelmed by the volume of information presented, which may lead to confusion rather than clarity.^31,32^ Finally, limited access to digital or printed PtDAs, due to socioeconomic factors, low health literacy, or mistrust in certain healthcare settings, can further restrict their use.^33–35^ Clinician-specific barriers that impact PtDA usage with patients are also important to consider.

Time constraints in the clinical encounter can prevent the clinician from using PtDAs and having SDM discussions.^36,37^ Access, including finding and retrieving the PtDA from the internet or an electronic platform may be a barrier for the clinician.^38^ Additional training may be required to learn how to effectively use the PtDA.^39^ Moreover, when cultural and language differences exist between the patient and clinician, using a PtDA that is not in the native language, or culturally appropriate, adds additional challenges to effective communication.^40,41^

### Patient portals as a platform for patient PtDA use

Patient portals are electronic, clinician-linked platforms that allow patients to view and use health information stored and managed within a healthcare organization.^42^ There is evidence that patients routinely use patient portals.^43^ While patient portals are not ubiquitous, 77% of patients in the US were offered access to their personal health information in 2024.^44^ In the same year, approximately 75% of patients managing a recent cancer diagnosis who had access to a patient portal logged in and used the portal at least once, compared to 65% of individuals nationally.^45^

Therefore, patient portals may be a viable option for delivering PtDAs to patients. Patient portals could be used to send PtDAs to patient/guardians before or after visits, or as part of a population health level campaign. Patient portal access is associated with a higher degree of patient engagement in health care,^46^ and use of patient portals continues to increase.^47,48^ Patient portals have been used to deliver education materials, including PtDAs^49^, with the aim of reducing access barriers so the patient and clinician can focus on communication. Patient portals could be used to increase the use of PtDA distribution and SDM at scale. However, it is important to note that accessing PtDAs via the patient portal could contribute to technological related barriers.^50^

This qualitative study aimed to elucidate patient’s access to and familiarity with PtDAs, as well as the barriers, needs, and requirements for adopting and using PtDAs received in various ways, including accessed via a patient portal. Focus groups (FG) were conducted across two academic medical centers to explore previous patient experiences with PtDAs, patient preferences for the receipt and use of PtDAs via a patient portal, and the contextual factors influencing the use of PtDAs to support SDM. Throughout this article, for clarity and brevity, we adopted terminology from the studies that are highlighted in the introduction. We use the term patient decision aid, and acronym PtDA, to be consistent with terminology routinely used by both the International Patient Decision Aid Standards (IPDAS)^51^ and the Cochran review.^13^

## Methods

We used the Consolidated Criteria for Reporting Qualitative Research (COREQ) for reporting guidelines (see Appendix 1).^52^ We used exploratory qualitative methodological orientation rooted in interpretivism^53,54^ and user-centered design^55–57^ (UCD). Focus groups were facilitated by BM, CR, and JF, while other study staff were present to take notes (EL). Patients were recruited at two US academic medical centers using purposive sampling as a technique to identify participants with PtDA expereince.^58,59^ The FG facilitators included BM who is male and holds a Ph.D., CR who is female and has a Master of Science in Nursing and is a Board Certified Registered Nurse, and JF who is female and is a human factors researcher; all three individuals work in health-related academic settings and have expertise in qualitative methods. Our research partners at the academic medical centers identified potential interview participants using existing networks such as patient panels or Community Advisory Boards (CAB).

Recommended individuals were recruited via email to gauge interest in participating in a FG. Eligibility requirements for the FG included individuals who were English speaking, 18+ years-of-age, and who self-reported having experience with using PtDAs. Participants were informed that the researchers had interest in their perspectives on PtDAs, patient portals, and receiving PtDAs via patient portals. Focus groups were conducted until we achieved thematic saturation.^60^ For this qualitative study, saturation was defined as not learning or hearing any new and substantive information from the patient participants, which was discussed by the qualitative team in regular consensus making meetings.

### Focus Group Guide and Data Collection

After a review of the literature to understand what was already known regarding patients’ use of patient-intended PtDAs are designed for patients/guardians to use independently, with reading, health, numeracy literacy at standard levels, language, schematic diagrams- as opposed to anatomical-detailed diagrams or images, and content appropriate for laypersons. Even with patients using patient-intended PtDA, it is still expected that patients will have further questions or need clarification from a clinician, and that the final decision may be made during a clinician-patient SDM discussion. Clinician-intended PtDA are designed to be used by medical professionals, who have the knowledge to use the PtDA to support a conversation with a patient/caregiver, and to translate the content to the patient’s education level, health literacy, language and other key factors. We (LS, AS, BM) developed an FG guide to gain insight on patient opinions regarding preferences, barriers, and facilitators to patients receiving and using PtDA obtained via patient portal. The FG guide was designed to allow participants to freely discuss emergent concepts and ideas that surfaced during cross-participant talk (see Appendix 2). The FG began with a short slide show to familiarize the patients with PtDAs some existing barriers for clinicians that try to use PtDAs with patients, and the different PtDAs formats, e.g., PDF, video, and web-based and EHR-integrated applications. We elicited patients’ experiences and perspectives using PtDAs and having SDM discussions with clinicians. This provided the FG facilitators with the necessary understanding of patient exposure to PtDAs to implement the remainder of the FG guide. The remainder of the questions focused on participants’ opinions regarding receiving and using PtDAs via a patient portal, their expectations for the patient portal, and any resulting follow-up communications, including SDM discussions. The discussion also covered barriers and proposed potential solutions to patient portal delivery and use of PtDAs. Each FG lasted one hour and was conducted on the Zoom™ ^61^ video conferencing platform. Sessions were recorded and transcribed verbatim for rapid analysis using a template and matrix approach.

### Rapid Template & Matrix Analytic Method

A four-step Rapid Qualitative Analysis(RQA)^62–65^ was conducted (BM, CR, EL, HB, JF) to systematically analyze and interpret the interview transcripts. First, a templated summary table was developed to ensure consistent data extraction, by domain, across all FGs transcripts. Domains included: 1) Previous PtDA Use, 2) Perspectives on PtDA Barriers and Facilitators, 3) Using PtDAs in My Existing Patient Portal, 4) Future PtDA Use and 5) Other (used to capture insights that did not fit into a predefined domain). Second, each FG transcript was summarized using this template, capturing key points and illustrative quotes by domain. Third, these summaries were compiled into a results summary matrix to facilitate cross-group comparison, reduce the data, and develop thematic synthesis by domain. Finally, the data was consolidated within a matrix to present summaries organized by topic domain, and developed overarching themes within, or across, domains.

#### Rapid Qualitative Analysis Step 1: Templated Summary

To quickly and efficiently achieve data reduction and thematic development, we (BM) created a template in which the 12 questions in the FG guide were grouped into five a priori domains^66–68^. We then iteratively revised the template as new emergent ideas came up in team discussion of how the template would be used by the qualitative team to extract relevant data.

#### Rapid Qualitative Analysis Step 2: FG Transcript Summarization

The qualitative team used the standardized template to summarize each unique FG transcript across five domains. Each domain summary was limited to four sentences, encouraging coders to prioritize key insights, and to reduce data. This technique, limiting the number of summary sentences, produces summary paragraphs that are more similar across coders and is important for the data reduction process. Two coders independently summarized each transcript; their summaries were then compared and refined through a consensus discussion. This process, in which discrepancies were discussed until agreement was reached,^69^ enhanced the rigor and reliability of the coding.^69–71^

#### Rapid Qualitative Analysis Step 3: Matrix Creation

A matrix was used to display each of the five domain summary paragraphs for each of the six FG transcripts. Next, two coders independently created a “summary of summaries” for each of the four domains, excluding the Other domain, which did not contain enough data to warrant a summary. Coders again aimed for four sentences, further aiding in the data reduction process and surfacing the most salient ideas into the summaries. Each pair of summaries for the domains were reviewed by the team for consensus and consolidated into one precise, and descriptive, overall domain summary paragraph.

#### Rapid Qualitative Analysis Step 4: Summary Consolidation and Theme Generation

Finally, we transferred each domain’s overall summary into a new matrix. These summaries were then uploaded to a Mural.co^72^ collaboration space to support discussion and theme development. Thematic analysis was guided by two principal factors: 1) the a priori domains organized data on topics central to the research question and narrowed the analytical focus, and 2) insights that emerged during the data reduction process, which helped distill the data into concise, descriptive themes.

All research procedures were reviewed and approved by the Colorado Multiple Institutional Review Board (COMIRB #22-1061).

## Results

Twenty-one individuals participated in six FGs across two academic medical centers between February and April 2024. Participant demographics are presented in Table 1. No participants dropped out of the study after being enrolled. Focus groups sessions were approximately 60 minutes. Data saturation was achieved after 6 FGs. All participants approached the discussion in the FG as a patient or family member, or both. The results were not returned to participants for member checking. Many participants baseline understanding of PtDAs confused PtDAs with general health education tools or questionnaires, although it became clear that all participants understood what a PtDA was during the FG discussion.

**Table 1.**
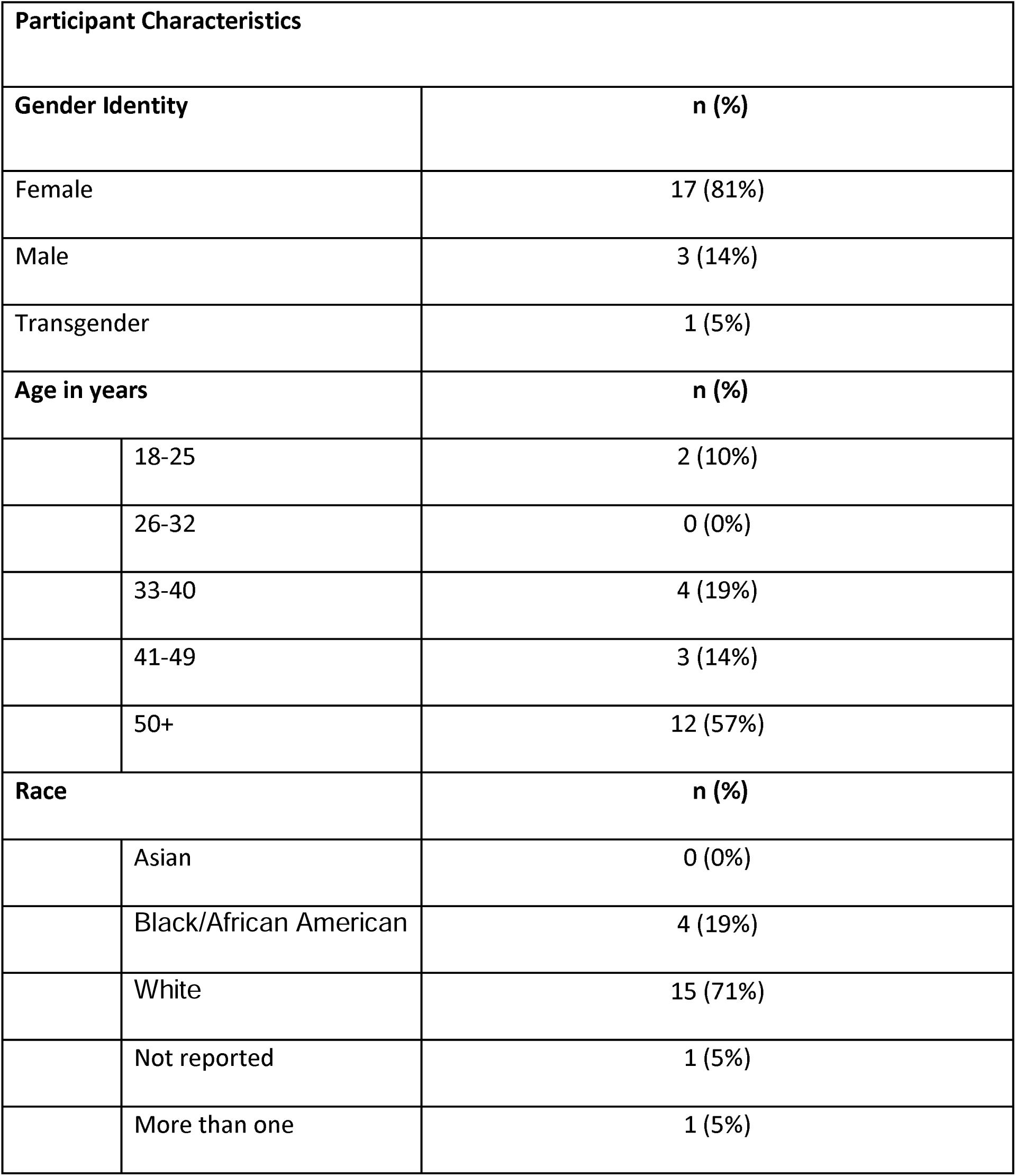
Focus Group Participant Characteristics (n=21)

### Theme 1: Patients had varied familiarity with PtDAs and often confused them with general health tools, but found value in SDM

Participants had varied experiences with the previous use of PtDAs with some having no previous experience, “Yes, well, first of all, I’ve never received a PtDA from a physician. I’ve never completed one. I’ve never been introduced to one” [FG001]. While experience with PtDAs was an intended eligibility requirement, some patients perceived other information received in a healthcare setting as examples of PtDAs. During deeper exploration of PtDAs during the FGs, it became clear there was confusion about the nature of what constituted a PtDA. As a result, no participants were required to leave the FG, and instead we provided continued explanation and clarification, as required during the FG sessions.

There were a few participants who had direct experience with PtDAs “Not in the digital format…I have had an experience with the process. I would say that I was given information about a variety of different possibilities with respect to treatment for me and pros and cons. And, and it was an engaging discussion in a structured fashion that I felt like I partnered with my doctor around. And so, it didn’t involve data points necessarily, but it did examine different possible outcomes of different treatment choices. And it was a really good conversation” [FG002]. Some of the participants had experience with PtDAs as a proxy, “I’m a caregiver for my father, and so we did one about his dialysis. He’s a dialysis patient, and so when he started doing dialysis, we did shared decision making with his doctor on whether he should do dialysis at home or whether going to a dialysis clinic” [FG005]. For those that did not have experience, many did not seem to understand the difference between PtDAs and other types of healthcare information. For instance, when asked about prior PtDA use, one participant provided an example of a “decision tree” document, “I had to complete a, it was a decision tree, and answer questions which led all the way down to medicine or surgery. And it was very helpful. I mean, it was wonderful. It was a great tool, and it was able to tell both of us what I needed” [FG006].

This participant described a PtDA but did not know to call the “decision tree” a PtDA. Other participants shared similar stories, but with different document formats, such as videos about what to expect at an upcoming procedure, pre-visit questionnaires, wearable monitoring devices, and apps to track physical therapy engagement.

In general, participants expressed that they benefitted from the SDM process, regardless of whether it involved the explicit use of a PtDA or not. For example, one participant shared what their clinician said, “…if you’re interested in looking at this different possibility that have some risk, and are new and cutting edge, but may well work on you. And it’s your choice. And then he [the clinician] went over everything. And so that was really fascinating to me, to look at all the different possibilities” [FG003].

### Theme 2: Patients perceived technology accessibility, PtDA credibility, and time as barriers potentially impeding PtDA use

Concerns across the FGs related to access and credibility. Our participants described being nervous about requirements to access technology, and the associated learning curve required to use the technology to get to the PtDAs. “Can I get access to it anytime? Do I have to log in and do this two-step access, or is it something like that, where he’s concerned (referring to a family member), he would just, say, ‘Forget it. I can’t get in. I’m not doing it’” [FG003]. Another participant directly addressed access, “I think it may be difficult if someone does not, you know, they may have a barrier when it comes to technology, possibly or lack of internet access, so they’re just unfamiliar with digital tools” [FG006].

Additionally, participants expressed that they would want assurance that the information in a PtDA was credible. Credibility included unbiased information that was consistently updated and would increase the potential for a beneficial outcome. Patients expressed concerns about the way PtDA were written, including health literacy levels, confusing medical jargon, and language barriers, “…it could be a culture, you know, sensitivity or differences in understanding health and medical decisions” [FG006]. The PtDA should be easy for patient-users to understand, use lay language and support SDM by providing information about how different options have impacted other patients, i.e., “I think if it’s something where you’re filling out information and it’s trying to make a decision for you, it’d be great if it gave you statistics for how that worked for other people” [FG006].

Patients identified having limited time with their clinician as the major barrier to PtDA use. “I have recently been in a situation where I had an appointment and was diagnosed with something very, very minor in comparison to lots of other things. But I didn’t feel like I got the information about that situation to my satisfaction, so I had to go home and Google it to find out more about it” [FG002]. Patients expressed concern that introducing the use of PtDAs would further burden conventional interactions with clinicians which are already time constrained. “I would not want this to cut into the examination or the discussion. And I know this is part of the discussion. It’s hard for me to think that a doctor can take all this time with a patient. Certainly, the most time that we have together is beneficial for me” [FG001]. Ultimately, patients want to feel encouraged to have such an open discussion with their clinician and confident that the PtDA presents accurate and relevant information.

### Theme 3: Patients valued flexible, user-friendly, and accessible PtDAs, particularly when introduced by clinicians and delivered via the patient portal, to support patient-led decisions

Participants emphasized the importance of PtDAs being valuable and that the discussion based on the PtDA should not be rushed, particularly for managing complex conditions. For complex decisions, previous familiarity of the PtDA by the clinician was deemed critical to be confident the PtDA was being used accurately, “I would have a greater sense of reliance on the basis of assuming that there is this understanding of, or prior exposure to, and ability to deal with the PtDA, that we’re engaged” [FG001].

Participants generally preferred to receive their PtDA electronically, but stated electronic materials might pose a barrier for older patients who are less comfortable with computers, harking to Theme 1. FG participants did assert they would prefer initial consultation in which an overview of the PtDA is provided by clinician before receiving any information through their portal. Participants also discussed preferences on how to receive a PtDA, with most participants preferring to receive it digitally, either through an email or text notification that would direct them to a message or link in the portal. Participants reported that short videos delivered through the patient portal explaining what patient decision aids are, demonstrating how they can be used to learn about diagnoses and the decision-making process, and encouraging clinician follow-up, would be helpful, “So long as there is a follow up from somebody in regard to anything that’s being asked by patients is a really important thing” [FG002].

Although participants expressed mixed views on the benefit of receiving a PtDA before a physician visit, many emphasized the importance of having access to PtDAs at essentially all times, i.e., before, during, and after the visit with the physician. Patients identified factors they wanted to know about PtDA before using it, including the purpose, goals, benefits, costs, supporting evidence or outcomes, and references. “I would want to know, you know, who’s the target audience? Is it a like, what are the groups or the levels or the backgrounds? What is the purpose for us to use this decision aid” [FG005].

In terms of decision making, these participants valued their clinician’s input, they stated the final decision should be their own, with the clinician offering guidance and openness to different choices. Participants wouldn’t mind having a SDM discussion supported by a PtDA with other trained healthcare team members, if these professionals are knowledgeable about the PtDAs and the decisions being made, “they better darn well know what they’re talking about” [FG004]. They also wouldn’t mind entering personal data into the PtDA, provided that was an easy process, but were concerned that inputting information themselves may introduce data entry errors and increasing patient burden for use, while also complicating the decision. “I think there’s a pro and con…inputting your own information. One risk there is, you might put it in wrong and get back a worse answer, but something I would kind of like is that you could also play around with your information” [FG001].

## Discussion

### Principal Findings

This qualitative study highlights the existing gap between the benefits and barriers of PtDAs, including SDM discussion supported by PtDAs, and their routine use in clinical practice. While PtDAs have been shown to improve patient knowledge, clarify values, and support SDM, the uptake of PtDAs is limited due to several patient-specific and clinician-specific barriers. Patients value PtDAs but face literacy, technological, and time-based barriers. Participants stressed the importance of using credible DA, having SDM discussion with clinicians who had expertise in the topic, and having adequate time for thorough non-rushed SDM discussion. This was important to them when decisions where more complex.

### Findings in Relation to Previous Research

Time constraints during visits compromise the clinician’s ability to introduce and explain PtDAs effectively, which is underscored by participant’s concern with the time allotted for patient-clinician appointments. The time issue may increase patient barriers to PtDA use that are multifaceted. Poor health literacy can prevent patients from understanding medical terminology, even when attempts are made a lower the grade-level of the language, and the implications of various options, limiting their ability to engage meaningfully in SDM. Emotional distress, particularly when facing serious diagnoses or irreversible choices, may lead to avoidance or disengagement from PtDAs.^73,74^ Historical and systemic distrust in the healthcare system, especially among marginalized populations,^29^ can reduce willingness to use PtDAs, based on the PtDAs lack of cultural and language sensitivities. Not having enough time to cover these topics can negatively impact patient uptake of PtDAs.

Beyond these well-documented barriers, our findings also underscore several underappreciated challenges. Patients may feel overwhelmed by the volume of information presented, leading to confusion rather than clarity.^31,32^ Socioeconomic factors, including lack of access to digital tools or printed materials, can restrict access to and usability of PtDAs. Patients often struggle to identify which PtDA is appropriate for their specific decision context, especially when tools are not clearly labeled or described.^75^ Therefore, as seen in this study, PtDAs that are introduced by the clinician are more trusted, which may lead to more patient uptake.

This qualitative study identified evidence that patient portals may be an acceptable, and even preferred, platform for delivering PtDAs. While participants were mixed on the timing of delivering PtDAs, emails or text messages were said to be helpful in locating and reviewing the content. Accessibility was raised as a barrier, indicating that there is a need for flexible delivery options, especially for older, less tech savvy patients. Our findings suggest that successful implementation of portal-based PtDAs may depend on integrating clinician endorsement, increasing patient trust in the PtDA, accessible educational supports with differing levels of health literacy options, all of which supports continued patient-clinician interaction, and that leaves the final decision up to the patient.

Ultimately, patients expressed concern with technical barriers to accessing PtDAs, particularly related to login and security features such as two-factor authentication. Delivering PtDAs through the EHR portal may facilitate access for patients who regularly use the portal, and 77% of patients in the US have been offered portal access in 2024.^44^ However, barriers persist after login. Participants emphasized that decision aids must be easy to use and accompanied by a clear explanation of why they were received. Patients made an important distinction between generic, or untailored messages, and PtDAs aids that were relevant to their specific situation and delivered at an appropriate point in the decision-making process, which could be before, during, or after an appointment with a clinician, depending on the circumstances. Participants indicated that provider communication was critical, including explaining the decision aid’s relevance and outlining the next steps in care.

### Implications

These findings provide guidance on future development of patient-centered SDM infrastructure and implementation strategies that aim to increase PtDA use. Health systems should prioritize clinician-supported delivery pathways, seamless integration of PtDAs into routine care and EHR workflows, and accessible materials that accommodate different levels of health literacy, technology comfort, and patient needs. If these factors are prioritized, implementation efforts may increase patient access, usage, and confidence in PtDAs.

Future work should evaluate implementation strategies that combine portal-based PtDAs that help coordinate delivery with clinician introduction and follow up. Research is also needed to assess whether health materials aimed at supporting patients, that are responsive to both health and digital literacy, have an impact on patient PtDA engagement, and ultimate decision-making. These inquires may help to identify gaps in meaningful PtDA access and use across a diverse patient spectrum.

### Limitations

This study has several limitations that should be considered. First, the sample was drawn from two academic medical centers and consisted primarily of patients already engaged with their healthcare systems, which may limit the generalizability of results to populations with lower healthcare engagement, or those in non-academic or rural settings. Participants were also required to speak English, and we sought participants who had previous experience with PtDAs, which may have excluded individuals with limited English proficiency, varied cultural backgrounds, or those unfamiliar with digital health tools, groups who may face distinct barriers to PtDA use. Participants also had to have access to the internet and be able to operate a computer or other device to use Zoom. Second, the FG context may have influenced participants’ willingness to share concerns or negative experiences about PtDAs, patient portals, or their interactions with clinicians. Social desirability bias may have contributed to more positive assessments of digital tools or clinician relationships. Finally, the study explored patients’ perspectives on hypothetically accessing PtDAs via patient portal rather than their experiences with an implemented system. Patient reflections were based on hypothetical interactions rather than real-world use. Future research that evaluates the portal in practice, across diverse clinical settings and patient populations, will be essential to validate and expand upon these findings.

## Conclusion

By articulating patient-derived needs and preferences for portal-based PtDA delivery, this study informs the design of SDM infrastructure that supports meaningful, patient-centered SDM in clinical practice. Addressing these barriers requires a multi-pronged approach. Health systems must invest in infrastructure that supports timely and tailored PtDA delivery, including accessible dissemination and culturally adapted materials. Timing is important in the introduction of PtDAs and could be determined based on the degree of complexity of the decision to be made. Future research should explore scalable strategies for improving PtDA accessibility, patient guidance, and integration into diverse care settings, including the patient portal, which has seen expanded use in recent years.

## Supporting information

Appendix 2 - SEDANS Patient Focus Group Guide

## Data Availability

All data produced in the present study are available upon reasonable request to the authors

## List of Abbreviations

CAB: Community Advisory Boards
DEAN: Decision Aid Navigator System
FG: Focus Group
PtDA: Patient Decision Aid
RQA: Rapid Qualitative Analysis
SDM: Shared decision-making

## Declarations

### Ethics approval and consent to participate

This study was reviewed and approved by the Colorado Multiple Institutional Review Board (COMIRB). The approved protocol included the conduct of FGs and qualitative analysis involving human subjects. All procedures were performed in accordance with institutional guidelines and relevant ethical standards for human subjects research. Informed consent was obtained from all participants prior to their participation in the FGs, including consent for recording and use of de-identified data in qualitative analyses.

### Consent for publication

Not applicable.

### Availability of data and materials

The datasets generated or analyzed during this study are available from the corresponding author on reasonable request.

### Author’s Contributions

BM, AS, and LMS designed the study. BM, CR, AN, EL, and HB, collected the data. BM, CR, AN, EL, and HB analyzed the data. BM, CR, AN, EL, HB, AS, and LMS drafted the manuscript. BM and LMS contributed to the interpretation of the results. All authors critically revised the manuscript and approved the final version.

## Acknowledgments

The authors would like to thank all the participants that participated in the discovery FGs to help us understand the usability patients expect and want from a medical patient portal.

## Clinical trial number

not applicable

## Appendix 1 COREQ – Consolidated Criteria for Reporting Qualitative Research

Patient Needs and Requirements for a Decision Aid Navigator System (DEAN) Patient Portal: A Qualitative Study
| Topic | Item Number | Guide Question/Description | Reported on Page No. |
| --- | --- | --- | --- |
| **Domain 1: Research team and reflexivity** |  |  |  |
| **Personal characteristics** |  |  |  |
| Interviewer/facilitator | 1 | Which author/s conducted the interview or focus group? | Pg. 7 |
| Credentials | 2 | What were the researcher credentials? e.g., PhD, MD | Pg. 7 |
| Occupation | 3 | What was their occupation at the time of the study? | Pg. 7 |
| Gender | 4 | Was the researcher male or female? | Pg. 7 |
| Experience and training | 5 | What experience or training did the researcher have? | Pg. 7 |
| **Relationship with participants** |  |  |  |
| Relationship established | 6 | Was a relationship established prior to study commencement? | N/A |
| Participant knowledge of the interviewer | 7 | What did participants know about the researcher? e.g., personal goals, reasons for doing the research | Pg. 8 |
| Interviewer characteristics | 8 | What characteristics were reported about the interviewer/facilitator? e.g., bias, assumptions, interests | Pg. 8 |
| **Domain 2: Study design** |  |  |  |
| **Theoretical framework** |  |  |  |
| Methodological orientation and theory | 9 | What methodological orientation underpinned the study? e.g., grounded theory, discourse analysis, ethnography, phenomenology, content analysis. | Pg.8 and Pg. 11 |
| **Participant selection** |  |  |  |
| Sampling | 10 | How were participants selected? | Pg. 7-8 |
| Method of approach | 11 | How were participants approached? | Pg. 8 |
| Sample size | 12 | How many participants were in the study? | Pg. 11 |
| Non-participation | 13 | How many people refused to participate or dropped out? Reasons? | Pg. 11 |
| **Setting** |  |  |  |
| Setting of data collection | 14 | Where was the data collected? | Pgs. 7, 9, 11 |
| Presence of non-participants | 15 | Was anyone else present besides the participants and researchers? | Pg. 7 |
| Description of sample | 16 | What are the important characteristics of the sample? e.g., demographic data, date | Pgs. 8, 11, 12 |
| **Data collection** |  |  |  |
| Interview guide | 17 | Were questions, prompts, guides provided by the authors? Was it pilot tested? | Pgs. 8 |
| Repeat interviews | 18 | Were repeat interviews carried out? If yes, how many? | N/A |
| Audio/visual recording | 19 | Did the research use audio or visual recording to collect the data? | Pg. 9 |
| Field notes | 20 | Were field notes made during and/or after the interview or focus group? | Pgs. 8- |
| Duration | 21 | What was the duration of the interviews or focus group? | Pg. 11 |
| Data saturation | 22 | Was data saturation discussed? | Pg. 8 and Pg. 11 |
| Transcripts returned | 23 | Were transcripts returned to participants for comment and/or correction? | Pg. 11 |
| Domain 3: Analysis and findings |  |  |  |
| Data analysis |  |  |  |
| Number of data coders | 24 | How many data coders coded the data? | Pg. 10 |
| Description of the coding tree | 25 | Did authors provide a description of the coding tree? | N/A |
| Derivation of themes | 26 | Were themes identified in advance or derived from the data? | Pgs. 2, 8, 9 and 11 |
| Software | 27 | What software, if applicable, was used to manage the data? |  |
| Participant checking | 28 | Did participants provide feedback on the findings? | N/A |
| **Reporting** |  |  |  |
| Quotations presented | 29 | Were participant quotations presented to illustrate the themes/findings? Was each quotation identified? | Pgs. 13- 17 |
| Data and findings consistent | 30 | Was there consistency between the data presented and the findings? | Pg. 18 |
| Clarity of major themes | 31 | Were major themes clearly presented in the findings? | Pgs. 2 |
| Clarity of minor themes | 32 | Is there a description of diverse cases or discussion of minor themes? | N/A |

Patient Needs and Requirements for a Decision Aid Navigator System (DEAN) Patient Portal: A Qualitative Study

Developed from: Tong A, Sainsbury P, Craig J. Consolidated criteria for reporting qualitative research (COREQ): a 32-item checklist for interviews and focus groups. International Journal for Quality in Health Care.2007. Volume 19, Number 6: pp. 349 – 357.

Once you have completed this checklist, please save a copy and upload it as part of your submission. DO NOT include this checklist as part of the main manuscript document. It must be uploaded as a separate file.

