## Appendix 2 - SEDANS Patient Focus Group Guide for "Patient Needs and Requirements for a Decision Aid Navigator System (DEAN) Patient Portal: A Qualitative Study"

SEDANS: Focus Group Guide

### Navigator Requirements: Patient & Proxies (CU only) needs

Purpose: To support patients in using PDAs that are relevant to their health and the decisions they make regarding their health, such as whether to have a certain procedure or whether to start or adhere to a certain medication.

Participants: Patients and caregivers that help make decisions in partnership with patients (proxies)

Method: In-person FG, with the ability to have some participants join via ZOOM (hybrid)

Introductory script:

Hello everyone! Welcome, and thank you for agreeing to be part of our discussion group today/tonight. Also, thank you for your interest in the Shared Decision Aid Navigator System (SEDANS) research project. Let me introduce our team: I’m ______ and I will be facilitating the discussion, and this is _________ and she/he will be observing, taking notes, and will likely also ask questions once we get into the discussion. We are from the University of Colorado Anschutz Medical Campus, and we are working with researchers from the School of Medicine who are interested in learning about what patients think about Patient Decision Aids (PDA) tools.

Do you have any questions about what a PDA is?

Specifically, we want to better understand your thoughts on how best to support the use of PDAs. We have asked you here today because you are or have been a patient. Your experience will help us design ways that patients, patient caregivers, and clinicians can use patient shared decision aid tools to have shared decision-making conversations. We have scheduled this focus group for 90 minutes. Participating in the discussion is up to you; you do not have to participate in the discussion if you do not want to do so.

Remember, the way we look at it, YOU are the experts here. We value your opinions. What you tell us during the discussion will help us design and develop a system to support PDA use and support patient decision-making.

Focus Group: ground rules:

Before we begin, let me mention a few things about how we usually conduct these groups:

1) I will be the facilitator for the group. My role is to ask the questions we have for the group and to encourage everyone to participate. I may ask you to explain more or to give an example while I am listening. Also, it’s my job to see that everyone has a chance to voice their opinions, as well as to keep us moving along so that we have time to discuss all the questions. So, at times, it might seem as though I am cutting you off, and this is not meant to be rude, but rather to make sure that we have time to have a complete discussion of each question.
2) It’s important that everyone hears this: there are no right or wrong answers. Each person’s experiences and opinions are valid, and we want to hear a wide range of opinions on the questions we’ll be asking. So, please speak up, whether you agree or disagree with what’s being said, and let us know what you think.
3) Sometimes participants bring up sensitive issues during these discussions, and we want to be sure that everyone agrees that anything of a personal nature that is mentioned in this room will NOT be repeated to others outside of this discussion group. Can I see a nod from everyone showing me that you agree with this confidentiality ground rule? (If anyone is not willing to give their consent to confidentiality, they may be excused from the group.) If a topic is uncomfortable to you, you do not have to participate or answer any questions.
4) One thing that is particularly important to note here is that this discussion group is NOT about us educating you on the use of PDAs. Today we’re going to focus the discussion on your views on using PDAs alone and/or with healthcare providers (ex: during a visit/encounter).
5) We need to emphasize that the discussion group is not an educational session where we give advice.
6) Let me tell you about our recording process. As you can see, we are using 2 digital recorders. We usually record these discussion groups because we want to capture everything that all of you say, and we simply can’t write fast enough to get it all down. We make every effort to protect your privacy by using only first names in the group discussion, and when we put together the results from all the groups, we won’t include any names at all. The recordings will be transcribed after the discussion. Once we have the written documents, the recordings are deleted.
7) It is very important that we speak one at a time so that the recorder picks up everything that is said. So, now that you know what our process is, is everyone OK with being recorded?
8) I’m also going to pass out note cards to each of you. I will collect these at the end of the session. If there is any feedback that you would prefer not to share out loud, that you prefer not to be recorded, or that you didn’t have a chance to say, please write it down on these cards and then give them to us after the discussion group is over.
9) We plan to finish our discussion by (time). When it is over, you will receive an electronic gift card as a thank-you to you for participating in our group today. Are there questions about any of this?
10) Please be sure to silence your phones if you have one with you.

Project overview:

This project aims to develop an electronic system that supports patients and providers finding and using Patient Decision Aids (PDAs) to make important medical decisions in partnerships. PDAs are tools that help patients and providers talk together about care options. PDAs help patients understand their choices about whether to have a specific treatment or screening. This includes knowing the pros and cons of each available option. PDAs also provide patients with the information needed to decide, based on their personal values and preferences, by exploring each option and how it relates to their values. For example, a PDA could help a pregnant woman decide whether to attempt a natural birth after having a cesarean. Research shows that PDAs lead to increased knowledge and a more accurate understanding of risk. When using a PDA, more patients feel comfortable with the decisions they make and the care they receive. The following questions do not have right or wrong answers. We would like your honest feedback and the preferences you have for using a PDA tool.

Guided Discussion:

- Please describe your previous use of PDAs.
  - Tell us about your experience…what was helpful, what was frustrating? What type of decision making (treatment, screening)?
- What stops you from using PDAs?
  - What would help address these barriers?
    - Plan to discuss each barrier…
- What information is important for you to know about each PDA you use?
- How do you decide to use, or not use, a PDA?
- Do you use MyHealthConnection, the patient portal?
  - If so, do you have any suggestions for how you would you want to see the PDAs organized in MyHealthConnection so you could find them? [show MHC portal screenshot]
    - What would help you find the PDAs in MyHealthConnection? How should the PDAs be categorized and/or organized in the patient portal?
    - Would you want to search through a list of PDAs to see if any were relevant to you or would you want your doctor to recommend them to you, or both?
    - Would you want to see PDAs that were not relevant to your health?
  - If not, how best should a clinician or practice staff give you information about a PDA?
    - e.g., Write it out on a printed after-visit summary, have a paper list of PDAs to give you after a visit, if possible, print out the entire PDA,
- How would you like to be notified about a PDA that is appropriate for learning/discussing a shared decision-making for your health condition?
  - MHC message
  - MHC dashboard
  - By the provider during a visit/encounter
- If a PDA was relevant to your health and the decisions you were making, how would you like to use the PDA?
  - Electronic Form
  - Paper form
  - Web application
  - Mobile application
- Can you imagine any trouble with using a PDA, recommended by your provider, as an electronic resource only? Would there be any reason you might like to download and print the PDAs?
  - Would you use a paper PDA...
  - ...during a visit with your provider?
  - ...to talk to your partner or spouse?
  - ...to share your thoughts with a proxy?
- Would you use a PDA in preparation for a doctor’s visit?
  - How about after a doctor’s visit?
  - Or would you only want to use it during a visit with your doctor?
    - Prompt: For example, if your doctor mentioned you starting a statin, would you use a suggested PDA after the visit (for learning more about the options/treatments) and get back to the doctor about your decision? Would you use it before the visit, in preparation for the visit and the discussion? How about if your doctor pulled it up on the computer screen—and you discussed it during the visit?
    - Prompt: Say it is a more complicated decision, like whether to have surgery or have a device implanted to correct your heart if it began beating in a way that is immediately deadly.
- If other members of the health care team, such as nurses, medical assistants, pharmacists, health coaches, were trained on the use of PDA and had the right background, how would you feel about using a PDA/having a discussion with them instead of your doctor?
  - Are there different considerations for the professional referenced above when using a PDA?
- How would you feel if you had to enter information such as your weight, age, lab values or other health conditions into a PDA to allow personalization of the PDA choices?
  - Would this be a barrier to using it?
  - Would it make you feel more confident in the information provided to you?
- If you have used a PDA in the past, would you like to go back and see, for example- cholesterol, prior results, review prior discussions with a physician?
  - If so, please explain your position.
- We would like to follow up on something earlier that you shared.
- Is there anything else you would like to tell us before we conclude?
